# Systematic analysis of homozygous autosomal copy number losses in exomes improves diagnostic yield and uncovers ultra-rare recessive disorders

**DOI:** 10.64898/2026.01.27.26344632

**Authors:** Ankur Chaurasia, Anju Shukla, Shruti Pande, Greeshma Purushothama, Akhil Kanathay Ashokan, Purvi Majethia, Namanpreet Kaur, Priyanka Upadhyai, Neha Quadri, Gandham SriLakshmi Bhavani, Dhanya Lakshmi Narayanan, Shalini S Nayak, Sheela Nampoothiri, Ataf H Sabir, Alaa A Mohammed, Sophie Shaw, Verity L. Hartill, Christopher M Watson, Colin A Johnson, Afrah Alshammari, Andrew E. Fry, James A. Poulter, William G. Newman, Paul R. Kasher, Siddharth Banka, Katta M. Girisha

## Abstract

Systematic analysis of copy number variants (CNVs) in large datasets is challenging and there are limited studies of homozygous copy number losses in rare disease exome datasets. Here we leveraged the genomic uniqueness and relative under-representation of the Indian population in the current public genomic databases and identified 42,386 possible homozygous losses (median count 20 per individual, range 0 – 55; median size 2.95 kb, range 99 bp – 4.76 Mb) in a heterogeneous cohort of 2,021 individuals with suspected Mendelian disorders, who had undergone exome sequencing using 12 different capture kits in a resource-limited setting. Employing a genomic position loss-count based approach, we filtered 1,224 rare homozygous loss calls in 718 individuals (median count 1 per individual, range 0 – 22; median size 3.49 kb, range 121 bp – 4.76 Mb) for further analysis, thus significantly reducing the analysis burden. Clinical correlation and validation of these rare calls enabled 10 new diagnoses in 240 unsolved individuals with at least one filtered rare homozygous loss call. This, led to nearly two-fold increase in diagnosis owing to homozygous deletions in our cohort. Further analysis of the data and identification of additional affected individuals through collaboration led to identification of biallelic *FILIP1* and *FAM177A1* variants as causes of a syndromic arthrogryposis and a neuromuscular disorder respectively. Both these conditions have been recently proven as ultra-rare recessive disorders, thus validating our approach. We also show that biallelic loss-of-function *TFCP2L1* variants cause chronic kidney disease and *VPS36* variants cause a severe recessive neurodevelopmental disorder characterised by microcephaly, motor delay, agenesis of the corpus callosum, cerebellar atrophy, seizures, hypotonia, spasticity and early death. Overall, these results demonstrate a scalable approach to screen homozygous losses for improving diagnostic yield and discovering disease-genes in large exome cohorts.

## Introduction

Even with exome and genome sequencing, genetic diagnosis is not achieved in the majority of patients with suspected Mendelian disorders^1^. This can be due to multiple reasons including difficulties in systematically incorporating information about copy number variants (CNVs), that are often complex and difficult to analyse. Short-read sequencing based CNV calling, for instance, suffers from high false-positive detection rates, making it difficult to distinguish true CNVs accurately^2^. Moreover, even when true CNVs are identified, their breakpoints may be imprecise or entirely unknown, a limitation particularly pronounced in exome sequencing data where breakpoints may not be covered by sequencing reads^3^. The lack of reliability and the heterogeneity in breakpoints and CNV size further complicate clustering or binning of variants for large-scale systematic analyses. Notably, exome sequencing is still preferred over genome sequencing particularly in resource-limited diagnostic settings. Collectively, these factors present significant obstacles to the reliable detection, filtering, and interpretation of the consequences of CNVs in large, short-read sequencing datasets.

Despite these challenges, recent studies have demonstrated the feasibility of using exome sequencing data for CNV analysis. For example, a heterogeneous exome cohort of 6,633 families (15,759 individuals) was analysed at the Broad Institute Center for Mendelian Genomics retrospectively using a read-depth analysis approach to detect rare causal CNVs in 171 families^4^. This approach was shown not only to improve diagnostic yield but also positioned exome sequencing as a possible higher-resolution alternative to microarrays while remaining more cost-effective than whole genome sequencing (WGS). Furthermore, large-scale reanalysis of 6,224 exomes of undiagnosed individuals from the Solve-RD project, integrating multiple structural variant (SV) detection methods based on paired-end read orientation, split-read signals, and read-depth data, uncovered additional 23 pathogenic SV including 15 CNVs, highlighting the advantages of using multiple complementary approaches in exome reanalysis projects^5^. These studies, along with several others^6–10^, demonstrate that incorporating CNV detection into large-scale exome analysis can lead to modest but meaningful improvements in diagnostic yield, ultimately helping end the diagnostic odyssey for families and reducing the need for additional testing.

Interpreting the consequences of complete gene knockouts resulting from homozygous losses may be relatively less challenging compared to heterozygous losses or other complex SVs such as gains, inversions, or translocations. One study leveraged this idea to analyse 11,754 rare disease parent-child trios and an additional 18,875 non-trio cases from the Genomics England 100,000 Genomes Project, focusing on an untargeted screening approach to detect inherited gene knockouts through homozygous deletions^11^. This study examined both known disease-associated and non-morbid genes, using cancer probands from the same cohort as controls. The findings demonstrated that while complete loss of morbid genes are rare, they can be reliably detected through adapted CNV analysis using cohort-wide data. Importantly, this approach led to 18 new diagnoses which had been missed by routine diagnostic pipelines. Similarly, in a dataset of 4,866 exomes at the Baylor–Hopkins Center for Mendelian Genomics, at least 15 new diagnoses due to rare homozygous deletions were detected by applying HMZDelFinder tool^12^. Both these studies underscore the potential of systematic homozygous loss screening in enhancing diagnostic outcomes. However, apart from these, there have been very few systematic studies investigating homozygous copy number losses in the context of rare diseases.

Endogamy is a common practice among Indian populations, contributing to higher inbreeding rates and unique genetic profiles with significant implications for genetic disease risk^13,14^. We previously generated a dataset of point variants in the Indian population using exome sequencing data from families with rare Mendelian disorders^15^. This dataset facilitated identification of novel disease genes and human knockouts. However, there has been no systematic large-scale study of CNVs in the Indian population despite its genomic uniqueness and relative underrepresentation in current public genomic databases.

In this study, we present a large-scale systematic analysis of homozygous autosomal coding copy number losses in 2,021 individuals from a consanguinity-enriched population in India. We developed a CNV calling pipeline, a filtering method, and prioritization strategies to efficiently identify and analyse 42,386 homozygous losses in exomes. We show that our genomic position-wise loss-count based filtering approach significantly reduces the analysis burden to a median of one rare homozygous loss per individual for clinical interpretation. New diagnoses found via this analysis led to a more than two-fold increase (from 9 to 19 diagnoses) in homozygous loss associated diagnostic yield of the cohort. We also provide evidence and independent replication to identify pathogenic variants in *VPS36*, *TFCP2L1*, *FILIP1*, and *FAM177A1* as causative for separate, severe recessive diseases.

## Materials and methods

### Exome data collection

This study was approved by the Kasturba Medical College and Kasturba Hospital Institutional Ethics Committee (IEC:856/2020). Informed consent was obtained from all participants for exome sequencing. Individuals affected by clinically suspected genetic disorders and their relatives who underwent exome testing at the Department of Medical Genetics, Kasturba Medical College, Manipal, India between 2011 and 2022 were studied retrospectively. DNA samples from these individuals were previously enriched and sequenced using twelve different exome capture kits. Sequencing reads were aligned to GRCh38 human reference assembly^16^ using bwa-mem2 (version 2.2.1; using default parameters)^17^. Binary alignment map (BAM) files were collected for CNV calling.

### Identification of homozygous loss calls

Based on the capture kit used for enrichment, CNVs were called in 12 independent batches using cn.mops^18^ (version 1.36.0) modules using modified parameters (I = c(0.025, 0.5, 1, 1.5, 2, 2.5, 3, 3.5, 4), classes = c("CN0", "CN1", "CN2", "CN3", "CN4", "CN5", "CN6", "CN7", "CN8"), priorImpact = 10, cyc = 30, parallel = 5, norm = 1, normType = "poisson", sizeFactor = "mean", normQu = 0.25, quSizeFactor = 0.75, upperThreshold = 0.85, lowerThreshold = −0.99, minWidth = 1, segAlgorithm = "fast", minReadCount = 1, useMedian = FALSE, returnPosterior = FALSE). Exonic coordinates for all possible transcripts were retrieved from the GENCODE gene annotation v38^19^, exons covered by the relevant capture kit were selected, and the reads mapping to the covered exons were counted. The read counts were then normalized, and an integer copy number change was estimated to call CNVs. Variants with zero copies (homozygous loss calls) from all batches were selected for subsequent analyses.

### Filtering of the called homozygous losses

Homozygous loss calls from all batches were mapped to the GRCh38 Human Reference Assembly by converting the chromosomal loss coordinates BED (browser extensible data) file into a BAM (binary alignment/map) file using BEDTools bedtobam with default tool parameters^20^. The count of mapped loss calls at each genomic position was then determined using Samtools depth with default tool parameters^21^. Genomic positions with loss-call count of fewer than six were identified and homozygous loss calls at these positions were filtered. A count of less than six was used, as we expected the same pathogenic homozygous position loss to be present in not more than 0.25% of the cohort (0.25% of 2,021 individuals is approximately 5.05).

### Annotation of the filtered homozygous loss calls

ClassifyCNV^22^ was used to annotate the filtered rare homozygous loss calls with genes. The association of the annotated genes with human diseases and their known inheritance patterns were ascertained using the Online Mendelian Inheritance in Man^23^ (OMIM) database. Calls that overlapped at least one known OMIM autosomal recessive (AR) disease gene and the calls that did not overlap any known OMIM AR disease genes were subsequently prioritised separately.

### Participants’ demographic data collection

Demographic data of the individuals with filtered rare homozygous loss calls were collected. Age, gender, and, where applicable, their relationship with the proband were recognized. Clinical phenotypes and diagnostic statuses were also noted for the probands and the affected family members. Details of the molecular diagnoses were recorded where applicable.

### Inspection of homozygous loss calls

Homozygous loss calls were manually inspected using Integrative Genomics Viewer (IGV)^24^. Exomes with loss calls were individually loaded with exomes of unaffected individuals for reference. Loss calls, for which high mapping quality reads (MAPQ > 30) were not found in the called region, when compared to the references, were deemed possibly true. Conversely, calls for which high mapping quality reads were found in the called region were deemed possibly false. Calls for which low mapping quality (MAPQ < 30) reads were found in the called region were deemed uncertain. Only the calls deemed possibly true were considered for further investigations.

### Prioritisation of calls with known OMIM AR disease genes

Homozygous loss calls overlapping known OMIM AR disease genes were first inspected using IGV, and possibly true calls were identified. Calls from unaffected individuals were excluded, and those from affected individuals were orthogonally tested using quantitative PCR (qPCR) from DNA extracted from blood. The predicted consequence of the loss on protein coding was evaluated for the principal transcript of the gene (the Matched Annotation from the NCBI and EMBL-EBI [MANE] representative transcript). For valid losses, phenotypes observed in the individuals were compared to the phenotypes reported in the OMIM database and the literature for the overlapping AR gene. If the clinical features of the affected individual matched the known phenotypes of the associated disorder, a diagnosis was reported to the family. Additionally, a survey of known variants and their types in these disease-causing genes was conducted using the Human Gene Mutation Database (HGMD®)^25^ and the published literature.

### Prioritisation of calls without any known OMIM AR disease genes

For homozygous loss calls without any known OMIM AR disease genes, calls from unaffected individuals and from previously diagnosed affected individuals were excluded. The remaining calls in the undiagnosed affected individuals were inspected using IGV to identify possibly true calls. Next, the dataset was examined to find loss calls that were more than 80% similar to the possibly true calls. Possibly true calls with similar loss calls in unaffected or phenotypically non-matching individuals were filtered out assuming similar losses may have similar phenotypic consequences. The remaining calls that had no similar loss calls in the dataset or had similar loss calls in individuals with overlapping phenotypes were selected. Genes in the selected calls were studied for the presence of homozygous predicted loss-of-function (pLoF) variants in control databases. Loss calls were excluded if the gene (or all the genes in a multi-gene call) had homozygous pLoF, single nucleotide, indel, or structural variants in gnomAD (v2.1, v3.1 and v4.1)^26^, human gene knockout dataset^27^, or our local unaffected individuals’ database. The remaining calls after exclusion were prioritized for orthogonal validation by qPCR using DNA sample extracted from blood. Genes in valid losses with no homozygous pLoF variants in the control datasets were regarded as candidate recessive disease genes.

### Candidate recessive disease genes study

Functional relevance of the gene to individuals’ phenotypes was studied. A gene was considered functionally associated if it had a relevant gene function and expression in the affected tissue or an overlapping animal model phenotype reported in the literature. The predicted impact of the loss on protein coding of the gene was evaluated for the principal transcript (MANE representative transcript). Gene expression was studied using the Genotype-Tissue Expression (GTEx) project dataset^28^. Animal models were searched using the International Mouse Phenotyping Consortium (IMPC) dataset^29^ and Zebrafish Information Network (ZFIN) database^30^. Additional individuals with biallelic variants in the candidate disease genes were searched in GeneMatcher^31^, the 100,000 Genomes Project (100KGP)^32^ tiering data, cohorts of other international collaborations, and literature. Variants were considered only when they were not present in homozygous state in gnomAD (v2.1, v3.1 and v4.1) and segregated with the phenotypes in the family.

### Validation of loss calls

Prioritized calls were validated and where available, segregation analysis in both the parents was performed using qPCR. The comparative quantification Ct (ΔΔCt) method^33^ was performed using the Applied Biosystems StepOne^TM^ Real-Time PCR System, Biosystems QuantStudio ^TM^ 5 Real-Time PCR System, and PowerUp^TM^ SYBR^TM^ Green PCR Master Mix (cat. no. A25741, ThermoFisher Scientific) according to the manufacturer’s protocol. Appropriate primer pairs were designed and used to test the amplification of exonic regions in the deleted genes. Data were analysed using StepOne Software v2.3 and QuantStudio Software v1.5.2. Relative DNA copy numbers were calculated using the expression 2×2^(−ΔΔCt)^.

### Fibroblast culture

Fibroblasts were cultured in Dulbecco’s Modified Eagle Media (DMEM, 4.5 g/L glucose, cat. no.10566016, Invitrogen) supplemented with 10% heat inactivated fetal bovine serum (FBS, cat. no. RM10409, HiMedia), 100 U/ml penicillin and 100 μg/ml streptomycin (cat. no. A018, HiMedia) and propagated in a humidified atmosphere (37°C, 5% CO_2_).

### cDNA synthesis and Sanger Sequencing

Total RNA was extracted from the fibroblast cells of F21 II.3 and an unrelated control using TRIzol™ (cat. no. 15596026, Invitrogen). Reverse transcription was performed using iScript™ Reverse Transcription Supermix (cat. no. 1725034, Biorad) according to the manufacturer’s protocol. PCR was performed using cDNA of F21 II.3 and control with the primer pair forward: ATCAACGAGACCCTGGTGAT and reverse: ACCTACAATTCCTACAGCCCT. The resulting amplicon of 601 bp encompassed the region between exon 1 and exon 6 of the *VPS36* gene. PCR amplicons were then purified (cat. no. FAGCK 001, FavorPrep™) and Sanger sequenced to assess the consequence of the homozygous loss.

### Immunoblotting

Cells were harvested using ice-cold radioimmunoprecipitation assay (RIPA) buffer (cat. no. 20-188, Merck Millipore) supplemented with mini protease inhibitor cocktail (cat. no. 11836153001, Roche) at 1X, on ice for 15 minutes. Cell debris were removed by centrifugation at 20,000g for 30 minutes at 4°C. Protein was quantified using QPRO-BCA Kit Standard (cat. no. PRTD1,0500, Cyanagen). Protein extracts (∼30 µg) were separated on sodium dodecyl sulfate-polyacrylamide gel electrophoresis (SDS-PAGE) and transferred to polyvinylidene fluoride (PVDF) membrane (cat. no. 1620177, Bio-Rad) using a semi-dry transfer system (cat. no. PB0010, Invitrogen). Membrane was blocked by incubating in TBS buffer (tris buffered saline with 0.1% Tween 20) supplemented with 5% blotting-grade blocker (cat. no. 1706404, Bio-Rad) for 1 hour at room temperature. The membrane was then incubated overnight at 4°C with primary antibodies (anti-Vps36 rabbit polyclonal, cat. no. 14262-1-AP, ThermoFisher Scientific; IB:1:100 and anti-GAPDH mouse monoclonal, cat. no. AM4300, ThermoFisher Scientific; IB: 1:2000) diluted in a blocking solution containing 5% bovine serum albumin (BSA) and TBS. PVDF membranes were then washed several times with 1X TBST and incubated with the Horse radish peroxidase (HRP) conjugated anti-rabbit and anti-mouse (cat. no. 1706515 and 1706516, Bio-Rad) secondary antibodies diluted at 1:10,000 for two hours at room temperature. Immunoreaction was detected using SuperSignal^TM^ West Pico Plus chemiluminescent substrate (cat. no. 34577, ThermoFisher Scientific). Immunoblots were digitally detected using iBright FL1500 imaging system (Invitrogen).

## Results

### Curation of rare homozygous loss calls from whole exome data

We assembled a cohort of 2,021 individuals, including affected probands and their relatives, from a population enriched for consanguinity and endogamy^15^ who had undergone exome sequencing at our centre from 2011 to 2022 using 12 different capture kits **(Figure 1a)**. The primary indications in the affected individuals included but were not limited to neurodevelopmental, musculoskeletal, immunodeficiency, and metabolic conditions. We developed a CNV calling pipeline and applied it to exomes in separate batches based on the capture kits used for enrichment. From this dataset we identified 42,386 possible homozygous losses (median count 20 per individual, range 0 – 55; median size 2.95 kb, range 99 bp – 4.76 Mb) **(Figure 1b)**.

**Figure 1:**
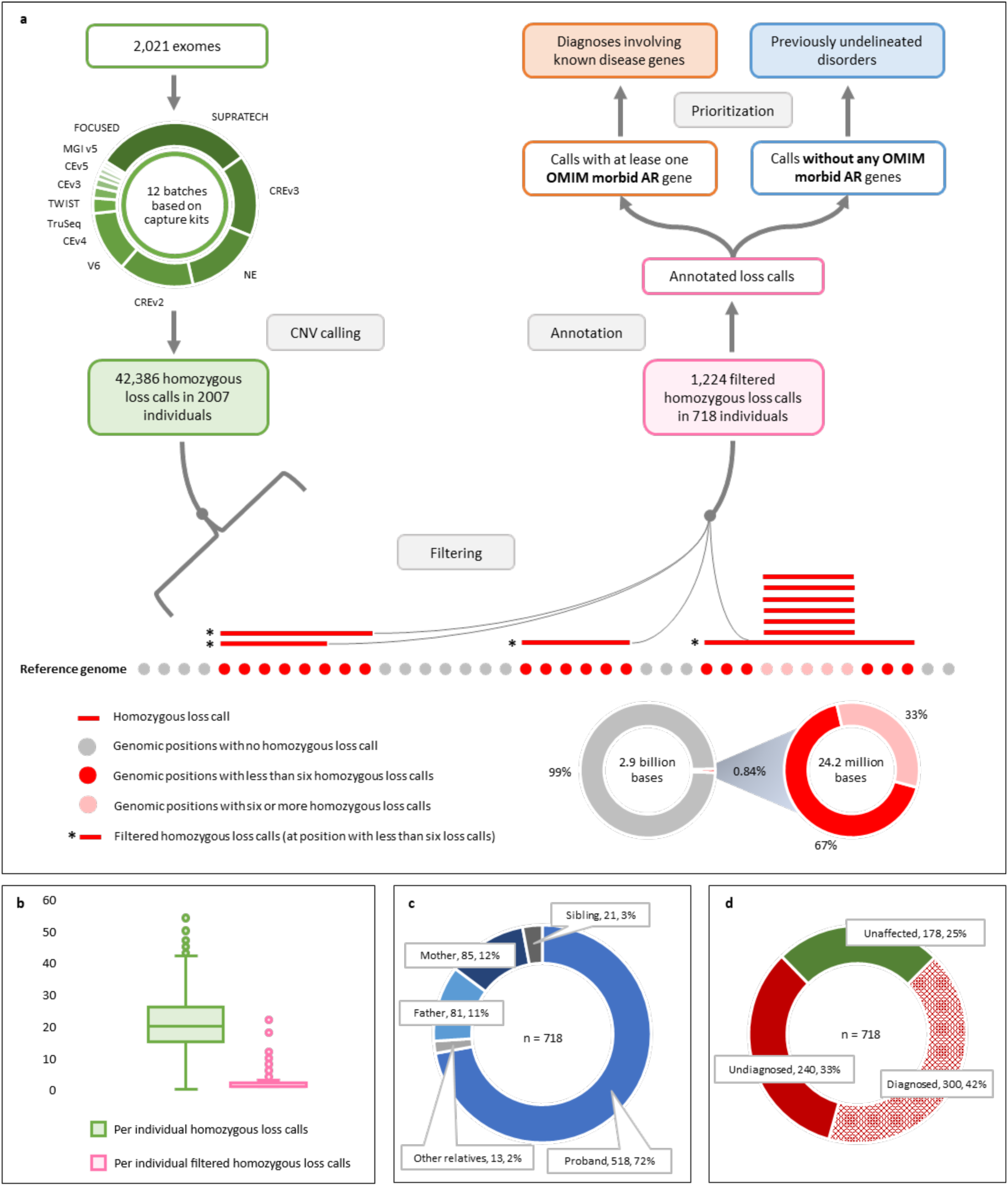
Homozygous loss calling, filtering, and annotation. (**a**) Analysis workflow. In total, 2,021 exomes from affected individuals and their relatives were collected for CNV calling. Exomes were processed in 12 separate batches based on the capture kits used. A total of 42,386 homozygous losses were called from these exomes. Calls were then mapped to the GRCh38 human reference assembly, and a position-wise count of the overlapping loss calls was determined. Positions with count of fewer than six were identified and calls at these positions were selected. The resulting 1,224 filtered rare homozygous loss calls from 718 individuals were annotated with genes, associated diseases, and inheritance patterns. Loss calls with or without known OMIM morbid autosomal recessive (AR) genes were prioritized separately using the attributes of individuals. Prioritized calls were then validated and clinically interpreted. (**b**) Per individual homozygous loss call counts before and after filtering. After CNV calling, a median of 20 homozygous losses was identified per individual (range, 0–55). Filtering reduced this number to a median of 1 filtered homozygous loss call per individual (range, 1–22). (**c**) relationship and (**d**) disease and diagnostic status of individuals with filtered rare homozygous loss calls.

From this dataset we wanted to identify calls that were rare and therefore more likely to be pathogenic. We defined rare calls as those with a relative semi-stringent internal frequency of five or less in our cohort. Usually in large datasets, the frequencies of similar overlapping CNVs are estimated by clustering them without considering the differences in their size and breakpoints^34^. However, we decided to take a genomic position-based approach to reduce the imprecise frequency estimation (i.e. due to dissimilar CNVs being mistakenly interpreted as being similar due to large overlap). For this, we mapped all the 42,386 loss calls to GRCh38 human reference assembly, counted loss calls at each base position, and determined positions that overlapped with five or fewer homozygous losses (16.3 million base positions). These positions were then used as anchors to filter the corresponding 1,224 rare homozygous loss calls in 718 individuals (median count 1 per individual, range 1 – 22; median size 3.49 kb, range 121 bp – 4.76 Mb) **(Figure 1b)**. 955 of 1,224 (78%) calls overlapped a single gene, 158 (13%) overlapped two genes, and 111 (9%) overlapped three or more genes. Only 97 (8%) loss calls overlapped a known OMIM AR disease gene. Analysis of the demographic data of 718 individuals with rare homozygous loss calls showed that 390 of the 718 individuals (54%) were male and 328 (46%) were female. Among the 718 individuals, 518 (72%) were probands, 166 (23%) were parents (81 fathers and 85 mothers), 21 (3%) were siblings, and 13 (2%) were other relatives **(Figure 1c)**. Of these individuals, 178 (25%) were unaffected, 300 (42%) had already received a molecular diagnosis, and 240 (33%) were affected and undiagnosed at the time of our analysis **(Figure 1d)**.

Overall, our filtering strategy significantly reduced the analysis burden (from median 20 to median 1) and led to the identification of rare homozygous loss calls from whole exome data.

### Prioritization of losses with morbid genes improves diagnostic yield

To prioritize potential diagnostic calls from the 1,224 rare homozygous loss calls, we selected those which overlapped one or more OMIM morbid AR genes. This revealed 97 calls from 74 individuals **(Figure 2)**. Following manual IGV inspection of all 97 calls we identified 25 possibly true calls in 24 individuals. Two calls detected in unaffected individuals were excluded from the analysis. Trio validation of the remaining calls performed by qPCR confirmed 19 out of the remaining 23 to be homozygous in affected individuals and heterozygous in parents. Three calls could not be assessed due to sample unavailability, and one was determined to be a false call.

**Figure 2:**
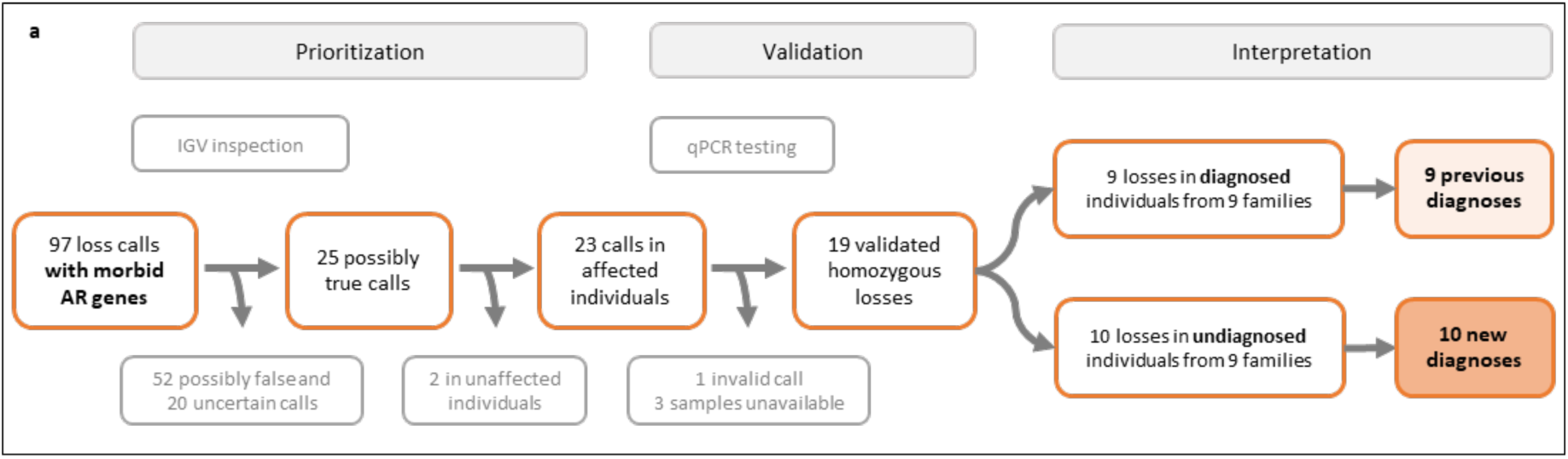
Prioritization, validation, and interpretation of filtered rare homozygous loss calls with a morbid AR gene. Results of prioritization, validation, and clinical interpretation. Calls overlapping known AR disease genes were inspected using IGV. Possibly true calls in the affected individuals were assessed for validity using qPCR. Confirmed homozygous losses were interpreted to establish diagnoses.

Firstly, we sought to examine the effectiveness of our filtering, prioritization and validation strategy. For this we compiled from our cohort a list of all homozygous deletions in AR genes that had been previously determined as causative variants through routine exome analysis. This search revealed nine such instances. These included seven partial single gene exonic deletions encompassing *CLCNKB* (AR Bartter syndrome, type 3, OMIM #607364), *CYP21A2* (AR congenital adrenal hyperplasia, due to 21-hydroxylase deficiency, OMIM #201910), *DOCK8* (AR hyper-IgE syndrome with recurrent infections, OMIM #243700), *FUCA1* (AR fucosidosis, OMIM #230000), *GM2A* (AR GM2-gangliosidosis, AB variant, OMIM #272750), *NPC2* (AR Niemann-Pick disease, type C2, OMIM #607625), and *OCLN* (AR pseudo-TORCH syndrome 1, OMIM #251290), and two contiguous multi-gene deletions, one in the 6p21.33 locus encompassing *PSORS1C1*, *C6orf15*, and the morbid *CDSN* (AR peeling skin syndrome 1, OMIM #270300) and the other in 16p13.3 region removing *HBZ*, *HBM*, *HBA2*, *HBA1*, and *HBQ1* causing hemoglobin Bart hydrops fetalis syndrome (OMIM #604131). Importantly, the 19 prioritized validated homozygous deletions included all nine previous diagnostic findings, thus validating our strategy.

The remaining 10 homozygous deletions were all found to be likely resulting in a partial or complete loss of function of the known OMIM-morbid gene **(Figure 2)**. We then performed reverse phenotyping^35^ of these 10 undiagnosed individuals (from 9 families). This revealed clinical features consistent with known disease descriptions, thus establishing new diagnoses in all 10 individuals. In brief, seven were partial single-gene exonic deletions, one each in *ADPRS* (AR neurodegeneration, childhood-onset, stress-induced, with variable ataxia and seizures, OMIM #618170)*, CNTNAP2* (AR Pitt-Hopkins like syndrome 1, OMIM #610042)*, DGAT1* (AR diarrhea 7, protein-losing enteropathy type, OMIM #615863)*, FANCA* (AR Fanconi anemia, complementation group A, OMIM #227650), and *PRKN* (AR juvenile Parkinson disease type 2, OMIM #600116), and two identical losses in *SLC13A5* (AR developmental and epileptic encephalopathy 25, with amelogenesis imperfecta, OMIM #615905) in a sib-pair. The remaining three losses resulted in contiguous multigene deletions, one at the 1p36.33 locus affecting *ATAD3B* and *ATAD3A* (AR pontocerebellar hypoplasia, hypotonia, and respiratory insufficiency syndrome, OMIM #618810), and two identical losses in the 2q13 locus affecting *MALL, NPHP1* (AR juvenile nephronophthisis 1, OMIM #256100), and *MTLN* in two unrelated individuals.

Next, we examined whether homozygous deletions were a recognized cause of the disease in the new diagnoses that we identified. Our investigation of the HGMD database and the literature revealed that homozygous exonic deletions have been documented in *CYP21A2*^36,37^, *DOCK8*^38,39^, *FUCA1*^40,41^, *GM2A*^42^, *NPC2*^43^, *OCLN*^44^, *CNTNAP2*^45–48^, *FANCA*^49,50^, and *PRKN*^51–54^, and full gene deletions have been reported in *CLCNKB*^55,56^ and *SLC13A5*^57^. Contiguous deletions in 6p21.33 region encompassing *C6orf15*, *PSORS1C1,* and *CDSN*^58^; 1p36.33 deletions encompassing *ATAD3B* and *ATAD3A*^59–63^; and 2q13 deletions encompassing *MALL*, *NPHP1*, and *MTLN*^64,65^ have also been reported. However, we found that exonic deletions in *ADPRS*, and *DGAT1* have not been previously reported, indicating that these deletions represent a new causative variant type in these genes.

Together, these results show that our focused analysis of homozygous CNV losses enabled 10 new diagnoses in 240 unsolved individuals (4.2%) with at least one filtered rare homozygous loss call. This led to a 2.1-fold (from 9 to 19) increase in diagnosis owing to homozygous deletions, thereby improving the diagnostic yield.

### Systematic analysis identifies candidate recessive disease genes

Next, we hypothesized that a systematic analysis of our data could lead to the identification of previously unrecognised recessive disorders. We, therefore, analysed the remaining 1,127 filtered rare homozygous loss calls from 681 individuals that did not overlap with any known AR disease genes **(Figure 3)**. Exclusion of calls based on individuals’ attributes, followed by IGV inspection, led to the identification of 96 possibly true calls in affected undiagnosed individuals. Phenotypic assessment-based prioritisation using our internal dataset and evaluation of affected genes in control databases (gnomAD^26^, human gene knockout dataset^27^, and our local unaffected individuals’ database) resulted in selection of 12 relevant loss calls that had at least one gene without homozygous pLoF variant in controls. Orthogonal trio validation performed using qPCR confirmed homozygous losses in the affected individuals and heterozygous carrier state of the respective parents in five families. These included seven partial single gene exonic deletions (1-4 deleted exons), with two identical losses in both *VPS36* and *TFCP2L1* in siblings of two families and one loss each in *FILIP1, FAM177A1*, and *PWWP3A*. These deletions were predicted to result in an absent, or truncated protein due to loss of transcription start site or frameshifts caused by the losses. To study the possible gene-disease associations, we sought additional similarly affected individuals who had biallelic deleterious variants in the five candidate genes. Individuals were searched via GeneMatcher^31^, 100,000 Genomes Project (100KGP)^32^ tiering data, international collaborations, and literature. We also studied the genes’ function, expression, and available animal models to understand their role in the conditions.

**Figure 3:**
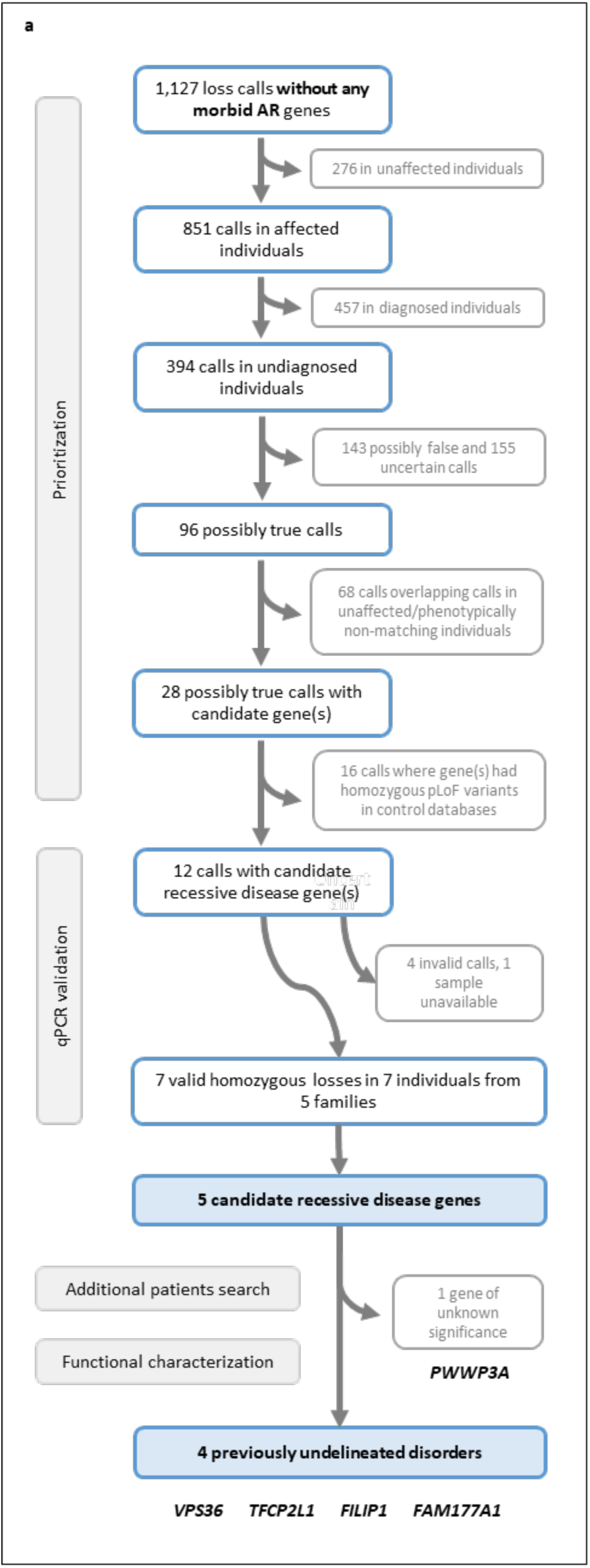
Prioritization of filtered rare homozygous loss calls without morbid AR genes and association study of the resulting candidate recessive disease genes. Results of prioritization, validation, and gene-disease association studies. Calls from affected undiagnosed individuals were inspected using IGV. Possibly true calls with no similar calls in unaffected or phenotypically non-matching individuals were selected. From these, calls overlapping genes with no homozygous pLoF variants in control databases were validated. Candidate recessive genes in confirmed homozygous losses were searched for additional similarly affected individuals with biallelic variants, and were studied for their function, expression in relevant tissues, and available animal models recapitulating the patient’s phenotype.

In Family 21 we identified a homozygous deletion encompassing exons 2 and 3 of *VPS36* (NM_016075.4) **(Figure 4)**. Additionally, through a GeneMatcher search, we identified three more families with homozygous *VPS36* variants and overlapping clinical presentations, along with a fourth family discovered through a literature search. Details of our *VPS36*-related work are provided in the next section.

**Figure 4:**
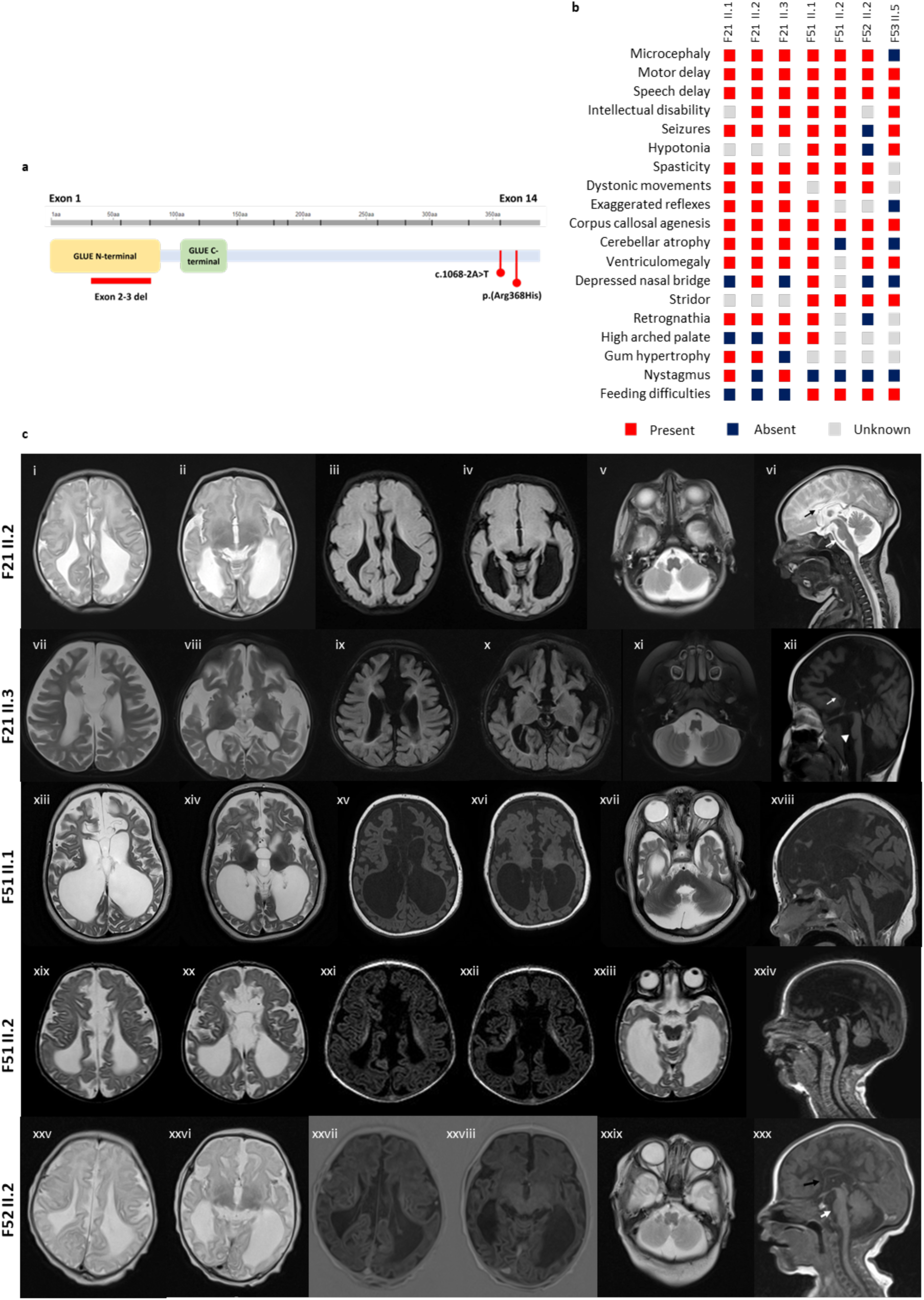
Genotypes, phenotypes, and brain MRI scans of individuals with bi-allelic VPS36 variants. (**a**) Schematic representation of the positions of identified variants across the VPS36 gene. *(b)* Key phenotypic characteristics of the seven individuals with homozygous VPS36 variants. (**c**) T1 and T2-weighted MRI scans of affected individuals’ brains, demonstrating key abnormalities. Axial and sagittal slices of the following individuals: F21 II.2 (first row; panels i–vi), F21 II.3 (second row; vii–xii), F51 II.1 (third row; xiii–xviii), F51 II.2 (fourth row; xix–xxiv), and F52 II.2 (fifth row; xxv–xxx) reveal pontocerebellar hypoplasia, posterior predominant ventriculomegaly due to agenesis of corpus callosum and white matter volume loss, and cerebral atrophy.

In Family 22, two siblings with homozygous deletions of exons 1 and 2 of the *TFCP2L1* gene (NM_014553.3) were identified to have chronic kidney disease, microcephaly, sensorineural hearing loss, and bilateral destruction of the femoral head, likely due to renal osteodystrophy. Additionally, another sibling with similar presentations and two spontaneous abortions of unknown cause were reported by the family. The homozygous loss identified in the two siblings is likely to result in a null allele due to the loss of transcription start site. Furthermore, through literature search, we identified another individual with a homozygous frameshift variant in *TFCP2L1* (NM_014553.3:c.869del:p.S290Ffs*50), who was affected with end-stage renal disease, congenital cataracts, hypotonia, seizures, and global developmental delay^66^. The variant was heterozygous in the parents and a non-identical unaffected twin and was predicted to result in premature termination of the protein prior to the sterile alpha motif (SAM)-like domain. *TFCP2L1* ortholog is expressed in mouse kidneys at both embryonic and adult stages and has been shown to induce growth and maturation of renal tubules in rat mesenchymal cells^66^. The orthologous transcription factor *Cp2l1* in mouse kidneys is also necessary for duct maturation^67^, indicating an important role of this gene in normal kidney function. Collectively, phenotypic convergence among the unrelated individuals identified in literature and our patients with chronic kidney disease, the deleterious effect of the variants on gene, and insights from animal studies together suggest that biallelic loss-of-function variants in *TFCP2L1* are associated with chronic kidney disease. The cause of other presentations in the two families is unclear and emphasizes on the need for a larger patient cohort to understand the full spectrum of phenotypes that may occur.

In Family 23, we identified an individual with homozygous *FILIP1* exon 3-6 deletion (NM_015687.5), multiple joint contractures, scoliosis, microcephaly, and facial dysmorphism. Our search identified seven individuals from four families with biallelic truncating variants in *FILIP1*. Two of these families, identified via GeneMatcher, had two distinct homozygous stop gains in three siblings (NM_015687.5:c.463G>T:p.E155*) and one child (NM_015687.5:c.2665C>T:p.R889*), respectively. A study detailing the genetic and clinical findings in these families, and in the individual we identified, was recently published seperately^68^. Another non-consanguineous British family was detected in the 100KGP with an individual with homozygous *FILIP1* stop gain variant (NM_015687.5:exon 5:c.2152C>T:p.R718*), presenting with joint contractures and dysmorphic features^69^. We also found a family in the literature with stop gain variants (NM_015687.5:exon 2:c.169C>T:p.R57*) for whom the two affected female siblings, homozygous for the variant, presented with hypotonia, joint contractures, motor delay, and delayed speech development^70^. *FILIP1* regulates filamin homeostasis through filamin A (FLNa) degradation^71^. It is required for the differentiation of cross-striated muscle cells during myogenesis^72^ and shows robust gene expression in both skeletal and smooth muscle^73^. It is also implicated in the regulation of neuronal cell positioning in the cortex^74^. Taken together, the phenotypic concurrence in the affected individuals, the predicted deleteriousness of all the identified variants, and the functional relevance of *FILIP1* in myogenesis, prove that biallelic loss-of-function variants in *FILIP1* are associated with a syndromic arthrogryposis.

In Family 24, we identified an individual with homozygous *FAM177A1* exon 5 loss (NM_173607.5) and spastic diplegia. We also found a mid-aged female with progressive neurological deterioration (detailed phenotype not available) and homozygous *FAM177A1* frameshift variant (NM_173607.5:c.218del:p.K73Rfs*93) in the 100KGP. Survey of the literature revealed nine additional individuals from four families with biallelic truncating *FAM177A1* variants and neurodevelopmental phenotypes^75,76^. Variants in the identified individuals and the variant in our individual with spastic diplegia were predicted to cause a loss of function as a result of protein truncation. *FAM177A1* is a ubiquitously expressed gene that encodes a Golgi complex protein of unknown function. It is thought that the gene is involved in intracellular transport of lipids between Golgi subcompartments by assisting VPS13B as a functional partner^77^. Collectively, the implicated functional role of *FAM177A1* in lipid transportation pathway, the disruption of which leads to similar phenotypes, the predicted deleteriousness of the identified variants, and the phenotypic concordance observed in individuals from unrelated families provide compelling evidence to associate biallelic loss-of-function variants in *FAM177A1* with neurodevelopmental disorder.

In Family 25, a homozygous deletion of exons 10-13 in the *PWWP3A* gene (NM_001369789.1) was detected in an individual exhibiting characteristics of spasticity and seizures. Two additional siblings in the family displayed similar phenotypes however, they were not available for genetic testing. The identified deletion is predicted to result in a frameshift, leading to the loss of coding sequence and consequently, gene function. No additional deleterious biallelic variants in *PWWP3A* were discovered in individuals from supplementary disease cohorts. PWWP domain-containing DNA repair factor 3A (PWWP3A) is a ubiquitously expressed nucleosome-binding protein that plays a significant role in the maintenance of the chromatin state. This protein operates as an accessory factor in the DNA damage response pathway, promoting chromatin modifications in response to DNA damage^78^. The *Pwwp3a* knock-out mouse model has been associated with hyperactivity and haematological phenotypes^29^; however, these phenotypes have not been evaluated in the reported individual. The contribution of *PWWP3A* to the observed clinical features remains undetermined.

Collectively, our systematic approach to prioritize rare homozygous loss calls, which did not overlap with any known AR disease genes, led to the identification of candidate autosomal recessive diseases which include *TFCP2L1*-linked renal tubulopathy, *FILIP1*-related arthrogryposis with microcephaly, and *FAM177A1*-associated motor dysfunction.

### Bi-allelic loss of function *VPS36* variants are associated with a neurodevelopmental syndrome

In Family 21, in three siblings with microcephaly, motor delay, agenesis of the corpus callosum, cerebellar atrophy, seizures, and spasticity, we identified a homozygous frameshift deletion encompassing exons 2 and 3 of *VPS36* (NM_016075.4) **(Figure 4)**. *VPS36* encodes EAP45, a component of the heterotetrameric endosomal sorting required for transport (ESCRT)-II subcomplex. Importantly, this complex includes *SNF8* encoded EAP30/VPS22^79^ **(Figure 5)** and the clinical features of affected individuals in family 21 showed a remarkable overlap with the known phenotype of disorders resulting from bi-allelic *SNF8* variants (Developmental and epileptic encephalopathy 115, OMIM #620783 and neurodevelopmental disorder plus optic atrophy, OMIM # 620784)^80^. Hence, we considered *VPS36* to be an excellent candidate gene for affected individuals in family 21.

**Figure 5:**
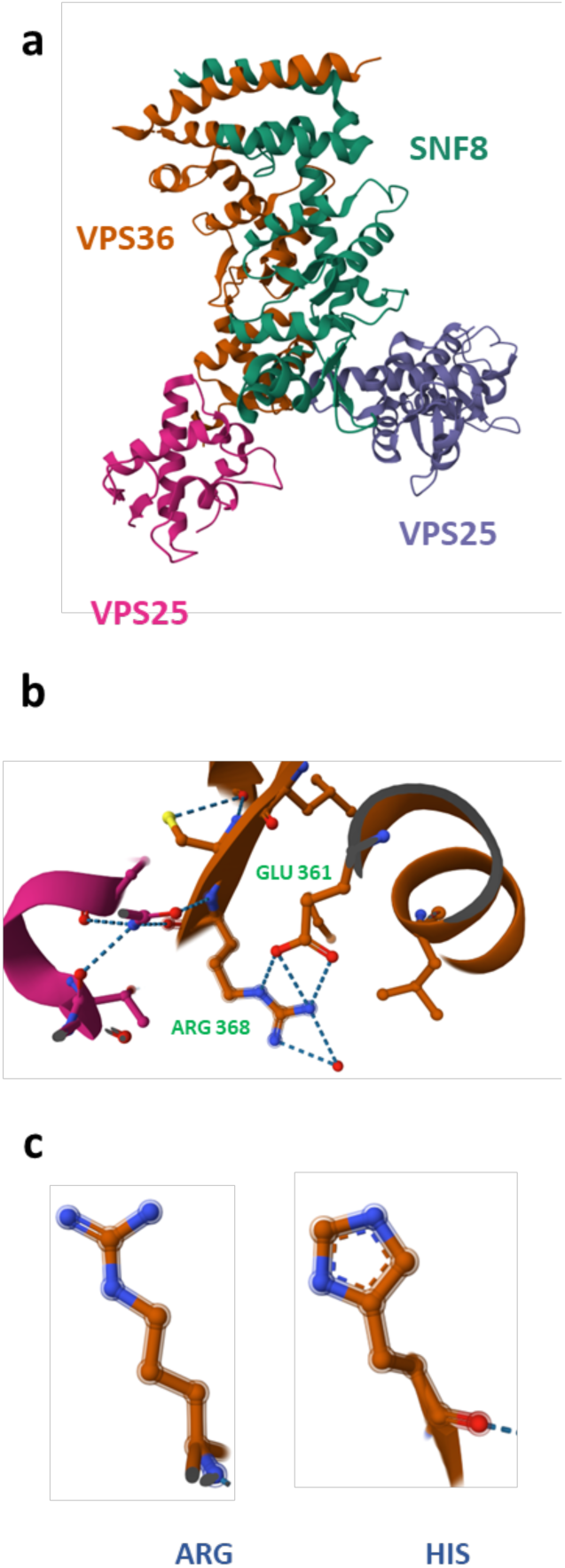
Structure of the ESCRT-II complex and the predicted impact of p.Arg368His substitution on VPS36 protein structure. (**a**) Crystal structure of the ESCRT-II complex (PDB ID: 3CUQ), showing its components: SNF8 (green), VPS36 (orange), and two VPS25 subunits (magenta and violet). (**b**) Detailed view of the interactions between the wild-type Arg368 residue and Glu361 within VPS36, highlighting intramolecular hydrogen bonds crucial for maintaining protein structural integrity. (**c**) Side-by-side comparison of the Arg and His side chains, illustrating that the p.Arg368His substitution likely disrupts native hydrogen bonding, thereby affecting the stability and overall structure of VPS36 and the ESCRT-II complex.

We next set out to identify additional individuals with rare bi-allelic *VPS36* variants. Literature search identified a homozygous *VPS36* likely splice-disrupting variant (NM_016075.4: intron 13: c.1068-2A>T; SpliceAI^81^ acceptor loss delta score of 0.97) reported in a diagnostic panels and exomes study from Saudi Arabia^82^ **(Figure 4)**. A gene-driven phenotype-agnostic GeneMatcher search revealed an identical homozygous *VPS36* missense variant (NM_016075.4:c.1103G>A:p.R368H) in two more families. The variant is rare in the heterozygous state (in eight of 806,918 individuals) and absent in the homozygous state in gnomAD. We found evidence of recent common ancestry between the two F51 and F53 families, but this haplotype was not shared with the F52 family. The variant has a high Combined Annotation Dependent Depletion (CADD) score of 33^83^ and predicted to disrupt native hydrogen bonding of residue R368 with E361, thereby affecting the structure and stability of EAP45 **(Figure 5)**.

Next, we examined the phenotypes of the additional individuals with rare homozygous *VPS36* variants. The individual identified via literature search was documented to have speech delay and intellectual disability, but no other phenotype details were available^82^. Family 51 consisted of two deceased male siblings, born to consanguineous parents, with microcephaly, corpus callosal agenesis, motor delay, intellectual disability, seizures, and hypotonia. Family 52 included a male child, born to consanguineous parents, with microcephaly, motor delay, dystonia, cerebellar atrophy, agenesis of the corpus callosum, and ventriculomegaly **(Figure 4)**. Notably, in Family 52, an older similarly affected female sibling had died early as a child. The *VPS36* variant in her could not be confirmed due to sample unavailability. However, previous SNP array data from her had shown a region of homozygosity, on chromosome 13, which includes *VPS36*. Family 53 consisted of two deceased males and one deceased female born to consanguineous parents. The female and one male had developmental delay, corpus callosum agenesis, ventriculomegaly, hypotonia and seizures (clinical in the brother and sub-clinical in the sister). The other male had corpus callosum abnormalities and ventriculomegaly but an additional skeletal dysplasia picture. Genetic testing confirmed a homozygous *VPS36* variant in the female. Testing of the males was not yet possible due to sample unavailability.

We then explored the functional consequences of the *VPS36* deletion in Family 21. Complementary DNA (cDNA) from fibroblasts of individual F21:II.3 demonstrated loss of the two exons in the *VPS36* transcript. Western blot analysis revealed a marked reduction in VPS36 protein levels in the fibroblast sample as compared to the control samples. It has been previously shown that presence of all ESCRT-II subunits is essential for its stability and if any one subunit is missing, the levels of the other subunits are reduced^80^.

Collectively, the rarity and deleterious nature of the *VPS36* variants, the striking phenotypic convergence observed in multiple affected individuals from unrelated families, and the functional relevance of the gene, and the significant clinical overlap with the known *SNF8*-related disorders suggest that bi-allelic *VPS36* variants cause a neurodevelopmental disorder.

## Discussion

Systematic analysis of large-scale genomic data can improve diagnostic yield and reveal previously unknown variants, genes and disorders^84^. It is challenging to implement read-depth based methods to identify CNVs in exome datasets generated from heterogenous cohorts. Identification and prioritisation of such CNVs at scale from exomes, especially those obtained from different platforms, batches and capture kits, remains challenging. Furthermore, lack of knowledge about the exact breakpoints of CNVs can make interpretation difficult. Comprehensive survey of homozygous CNV losses in large disease datasets has been attempted in a limited number of studies^4,5,12,85^.

In this study we successfully devised and implemented a strategy to re-analyse homozygous deletions from a heterogeneously generated historical dataset in a rare disease cohort enriched for consanguinity. This was achieved by calling CNVs separately in capture kit dependant batches followed by integrating them into a single database. Most large scale CNV studies from WES or WGS utilise overlaps of CNVs to classify them as similar and to determine their allele frequency^4,34^. However, this approach overlooks the fact that CNVs sharing a common overlap can still vary in size and breakpoints, potentially disrupting distinct genomic regions. Consequently, this method may erroneously exclude CNVs affecting pathogenic loci if their allele frequency is inflated due to overlapping yet non-identical CNVs. To address this limitation, we prioritised rare CNVs based on their frequency at the individual nucleotide level, using a genomic position-wise loss-count based filtering strategy. This refinement reduced the analysis burden by 97.1%, thus considerably decreasing the number of losses requiring expert evaluation. Notably, none of the previously known homozygous loss-associated diagnoses were missed, thus reducing the chances that our strategy resulted in discarding of some relevant deletions. This approach revealed new diagnoses and led to the identification of candidate disease genes. Overall, we identified 19 individuals in our cohort whose diagnoses could be attributed to a homozygous CNV loss. Thus, this analysis increased the number of individuals with diseases due to homozygous deletion by 2-fold and thus significantly increased the diagnostic yield. Taking together seven more individuals with new disease gene associated presentations, homozygous losses accounted for phenotypes in 26 of the 240 (10.8%) unsolved individuals in our cohort who had at least one rare homozygous loss call.

However, there are several limitations of the approach. Firstly, we experienced a high false detection rate. This is possibly due to the highly heterogeneous sequencing data collected over a period of 12 years from various service providers. Although we excluded uncertain calls from our analysis, they can still represent valid biological losses. Furthermore, in our analysis, we focused only on MANE Select canonical transcripts^86^. While MANE transcripts offer consistency, analysing alternative isoforms would be crucial for comprehending the complete functional consequences of the losses and the underlying disease mechanisms. Moreover, exact breakpoints were not assessed in most cases, and the possibility of the loss being part of complex rearrangements cannot be ruled out. To prioritize loss calls in our analysis of new candidate recessive disease genes, we excluded calls from previously diagnosed individuals. Although these individuals have a confirmed diagnosis, they may still carry a pathogenic loss in a gene not yet linked to disease. This could contribute to a blended phenotype or a presentation closely resembling the known disease associated with the diagnosed gene, raising the possibility of a dual diagnosis. Moreover, we did not explore the functional consequences of the losses identified in unaffected individuals. Such losses could represent natural human knockouts that do not lead to a clinical phenotype, offering broader implications for understanding the functional roles of such genes^27^. Potentially, supplementing our approach with paired-end and split read-based structural variant (SV) detection tools ^5,85^ and long-read sequencing technologies may help to address some of these issues at scale.

Our work provides evidence to support the existence of *FILIP1*-related arthrogryposis with microcephaly, *TFCP2L1*-related renal tubulopathy, *FAM177A1*-related motor dysfunction and *VPS36-*related neurodevelopmental disorder. *FILIP1*-related congenital neuromuscular disorder with dysmorphic facies (OMIM #620775) is now indexed in OMIM and has been shown to be characterized by impaired skeletal muscle development, usually resulting in hypotonia and secondary joint contractures, and dysmorphic facial features. Overall, this validates our approach. With regards to *TFCP2L1*-related renal tubulopathy, it is noteworthy that the affected individuals also exhibit other phenotypes (microcephaly, sensorineural hearing loss, congenital cataracts, hypotonia, seizures, and global developmental delay) that are not directly attributable to the renal dysfunction. This could be due to the known role of *TFCP2L1* beyond kidney development and function^87–89^. *FAM177A1*-related motor dysfunction seems to be characterized by global developmental delay, seizures, behavioral abnormalities, hypotonia, and gait disturbance. Although the cellular function of FAM177A1 is not fully understood, it has been observed to localize in the Golgi subcompartments and may be involved in the intracellular lipid and protein trafficking^77^. Identification of more individuals with these conditions will be important for better characterisation of the disorders.

We also provide genetic, clinical and functional evidence to support a relationship between homozygous *VPS36* variants and a neurodevelopmental disorder characterised by microcephaly, motor delay, agenesis of the corpus callosum, cerebellar atrophy, seizures, hypotonia, spasticity, and early death. The studies performed using fibroblast cells of F21 II.3 confirmed the loss of exons 2 and 3 in the *VPS36* transcript. Although the precise mechanism by which this deletion causes disease is unclear, it is likely that the deletion impacts the stability of the ESCRT-II complex, of which VPS36 is a subunit, thereby disrupting the function of this highly conserved assembly. This ESCRT system comprises three distinct multi-subunit complexes, ESCRT-I, ESCRT-II, and ESCRT-III, and plays a critical role in various cellular processes, including cytokinesis, endosome maturation, autophagy, and membrane repair^90^. In humans, the ESCRT-II complex includes one subunit each of VPS36 (EAP45) and SNF8 (EAP30/VPS22) and two subunits of VPS25 (EAP20)^79^. Mouse knockout models of *Vps36* and other ESCRT subunits result in early embryonic or preweaning lethality^80,91^. Another subunit of the ESCRT-II complex, SNF8, has been associated with remarkably similar phenotypes, and biallelic variants in *SNF8* have been shown to reduce the protein levels of other ESCRT-II subunits (VPS36 and VPS25)^80^. Furthermore, several other genes encoding subunits of the ESCRT (consisting of ESCRT I to III subcomplexes) machinery have been implicated in a range of neurodevelopmental disorders in humans, such as AR spastic paraplegia 53 (*VPS37A*; OMIM #614898), AD spastic paraplegia 80 (*UBAP1*; OMIM #618418), AR pontocerebellar hypoplasia type 8 (*CHMP1A*; OMIM #614961), AD frontotemporal dementia and/or amyotrophic lateral sclerosis 7 (*CHMP2B*; OMIM #600795), and AD CIMDAG syndrome (*VPS4A*; OMIM #619273). This suggests that the ESCRT machinery plays an important role in normal development and its impairment can lead to a spectrum of neurodevelopmental conditions in humans.

In summary, employing filtering strategies for CNV screening can streamline the analysis process. By integrating these techniques with in silico phenotype matching, it is possible to scale up the analysis and make it feasible for large cohorts. This can also make periodic re-analysis possible which has been proven to improve the diagnostic yield of the cohorts and is becoming a common practice across both clinical and research settings. Although we focussed on identification of disease-causing homozygous deletions, it is possible to customise this approach to identify heterozygous CNVs or to analyse copy number gains from large genomic datasets. To conclude, we developed a CNV calling pipeline to identify homozygous losses from exomes and applied filtering and prioritisation strategies to facilitate diagnostic screening and rare disease discovery in a cohort of 2,021 individuals. Our methods increased homozygous loss associated diagnoses by more than two fold and defined four new recessive diseases caused by biallelic variants in *VPS36*, *TFCP2L1*, *FILIP1,* and *FAM177A1*. To better understand the clinical variability of these disorders and establish more accurate genotype-phenotype correlations, it will be essential to identify additional individuals and perform further experimental research to investigate the functional effects of these pathogenic variants in the newly discovered disease genes. This information will help to develop effective disease management strategies and potential therapeutic targets that may slow or prevent disease progression.

## Data and code availability

The data and code used in this study are available from the corresponding author upon reasonable request.

## Author contributions

Funding was acquired by SB, CAJ and KMG. The study was conceptualized by AC, SB, and KMG. Methodology was developed by AC, AS, NK, PU, SSN, SS, VLH, CMW, EH, AEF, JAP, PRK, SB, and KMG. Investigations were conducted by all authors. Data curation and analyses were performed by AC, AS, SP, GSLB, PU, NQ, PM, AA, SB, and KMG. Project administration and supervision were provided by PRK, WGN, CAJ, SB, and KMG. The original manuscript draft was prepared by AC, SB and KMG which was reviewed and approved by all authors.

## Acknowledgements

The authors express gratitude to all the patients and their family members who consented to sequencing their exomes at participating centers.

## Web resources

OMIM, https://www.omim.org/

gnomAD, https://gnomad.broadinstitute.org/

Genotype-Tissue Expression (GTEx) Portal, https://www.gtexportal.org/home/

GeneMatcher, https://genematcher.org/

The Human Gene Mutation Database (HGMD®), https://www.hgmd.cf.ac.uk/ac/index.php

The International Mouse Phenotyping Consortium (IMPC), https://www.mousephenotype.org/

The Zebrafish Information Network (ZFIN), https://zfin.org/

